# Hepatic steatosis and steatohepatitis: a functional meta-analysis of sex-based differences in transcriptomic studies

**DOI:** 10.1101/2020.06.03.20118570

**Authors:** José F. Català-Senent, Marta R. Hidalgo, Marina Berenguer, Gopanandan Parthasarathy, Harmeet Malhi, Pablo Malmierca-Merlo, María de la Iglesia-Vayá, Francisco García-García

**Affiliations:** Bioinformatics and Biostatistics Unit, Principe Felipe Research Center, Valencia, Spain; Spanish National Bioinformatics Institute, ELIXIR-Spain (INB, ELIXIR-ES); Liver Transplantation and Hepatology Unit, Hospital Universitario y Politécnico La Fe, València, Spain; Grupo de hepatología, Cirugía HBP y Trasplantes, Instituto de Investigación Sanitaria La Fe, València, Spain; CIBERehd, Centro de Investigación Biomédica en Red en Enfermedades Hepáticas y Digestivas, Instituto de Salud Carlos III, Madrid, Spain; Department of Medicine, Universitat de València, València, Spain; Division of Gastroenterology and Hepatology, Mayo Clinic, Rochester, MN, USA; Atos Research Innovation (ARI), Spain; Biomedical Imaging Unit FISABIO-CIPF. Fundación para el Fomento de la Investigación Sanitario y Biomédica de la Comunidad Valenciana. Valencia, Spain

**Keywords:** Non-alcoholic Fatty Liver Disease, Sex Characteristics, Precision Medicine

## Abstract

**Background & aims:** Sex differences in non-alcoholic fatty liver disease (NAFLD) are well known and yet, most of the studies available in the literature do not include this factor when analysing the data. Here we present a functional meta-analysis of NAFLD studies to detect sex-related alterations of molecular mechanisms, between stages of the disease.

**Methods:** We systematically reviewed the Gene Expression Omnibus database functional Gene Ontology terms detailed in transcriptomic studies, following the PRISMA statement guidelines. For each study, we compared steatosis (NAFL) and steatohepatitis (NASH), in premenopausal women and men using a dual strategy: a gene-set analysis and a pathway activity analysis. The functional results of all the studies were integrated in a meta-analysis.

**Results:** A total of 114 abstracts were reviewed and 7 studies, which included 323 eligible patients, were finally analysed. The meta-analyses highlighted significant functions in both sexes. In premenopausal women, the overrepresented functions referred to DNA regulation, vinculin binding, IL-2 responses, negative regulation of neuronal death, and the transport of ions and cations. In men, they referred to the negative regulation of IL-6 and the establishment of planar polarity involved in neural tube closure.

**Conclusions:** Our meta-analysis of this transcriptomic data provides a powerful approach to identify sex differences in NAFLD, when comparing NAFL and NASH. We detected relevant biological functions and molecular terms that were affected differently between premenopausal women and men. Changes in the immune responsiveness between men and women with NAFLD suggested that women have a more immune tolerant milieu while men have an impaired liver regenerative response.

**Synopsis:** Progression from NAFL to NASH differently affects cellular functions in women and men. Here we systematically reviewed publicly available transcriptomic data and then performed a meta-analysis to find these affected functions. Thus, we identified 13 biological functions implicated in the progression of NAFLD that were differentially affected by sex.

**Graphical abstract:** 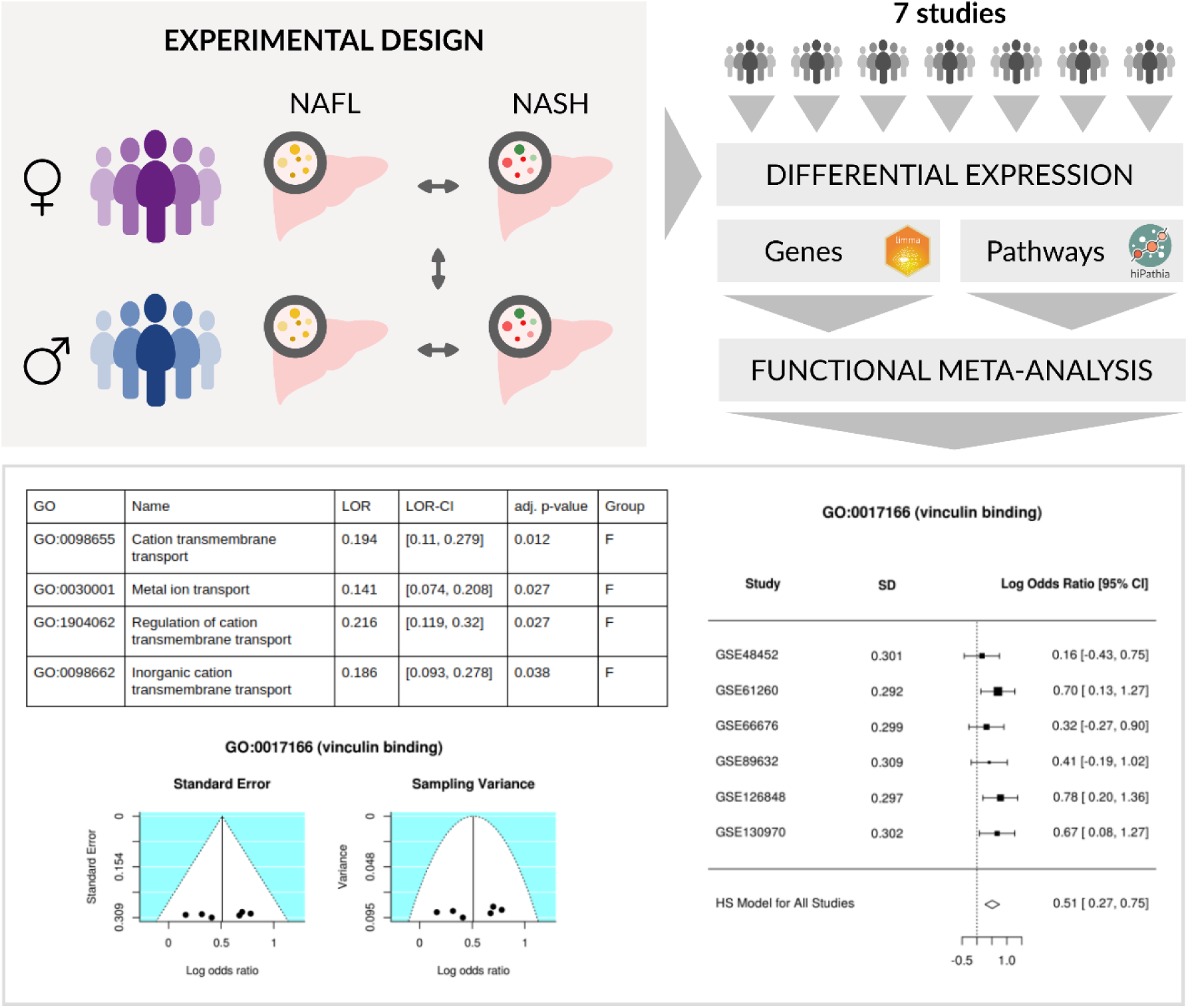

## Introduction

Non-alcoholic fatty liver disease (NAFLD) encompasses a spectrum of liver disorders ranging from fat accumulation in hepatocytes (NAFL) to non-alcoholic steatohepatitis (NASH), which can all potentially lead to cirrhosis or liver cancer. Interest in this disease has increased in recent years because of its worldwide impact on health, given that it is now the most common liver disease in developed countries ^1^. It is estimated that the worldwide prevalence of this disease is around 25%, although it is more common in South America and the Middle East and less prevalent in Africa ^2^.

Sex also affects the distribution of this pathology, with current studies showing a higher prevalence in men and postmenopausal rather than premenopausal women ^3^. Indeed, the liver shows the second largest amount of sexual dimorphism in humans. Both physiological and pathological hepatic processes, such as the detoxifying metabolism of cholesterol, as well as the prevalence of hepatic diseases differ between men and women ^4^. Hence, the progression of NAFLD and the effect of treatments are also likely affected by patient sex ^5^. Nonetheless, most studies on this disease do not consider metabolic differences between the sexes ^6^.

Although the presence of NAFLD can be strongly suspected based on imaging results, clinical analyses, and the presence of comorbidities, histological analysis is the only definitive way to diagnose it ^7^. Because of its invasive nature, the fact that a liver biopsy is the only completely reliable technique to diagnose NAFLD is a limitation when conducting studies in human patients.

The transition of NAFLD stages is different in each patient, and can have a considerable impact on the clinical consequences to the individual. Patients in the initial stage of the disease (NAFL) have a low risk of adverse outcomes ^8^, but its progression to NASH increases the possibility of clinical complications, both liver-related and otherwise.

In this context, we carried out an exhaustive review and selection of transcriptomic studies which had deposited data in the Gene Expression Omnibus (GEO) datasets database ^9^, to guarantee the eligibility of the selected studies. We used these to clarify the molecular basis of the differences between NAFL and NASH, in premenopausal women and in men. Sex-based differences may have important clinical implications to patients. This knowledge could potentially favour a personalised approach to this disease in the future.

## Results

### Systematic review and exploratory analysis

As shown in the PRISMA flow diagram (Figure 1), our systematic review yielded 114 non-duplicated studies. After applying the exclusion criteria described above, a total of 7 studies were included in this work. Studies were excluded for the following reasons: studies not in humans or unrelated to NAFLD (*n* = 55), in vitro studies (*n* = 15), lack of information on sex or inclusion of only one sex (*n* = 24), no patients had the NAFL and NASH disease stages (*n* = 12), and diagnoses not based on histology (*n* = 1). The strict level of these inclusion and exclusion criteria allowed us to select a set of comparable studies to ensure the reliability of the subsequent analysis strategies.

**Fig. 1.**
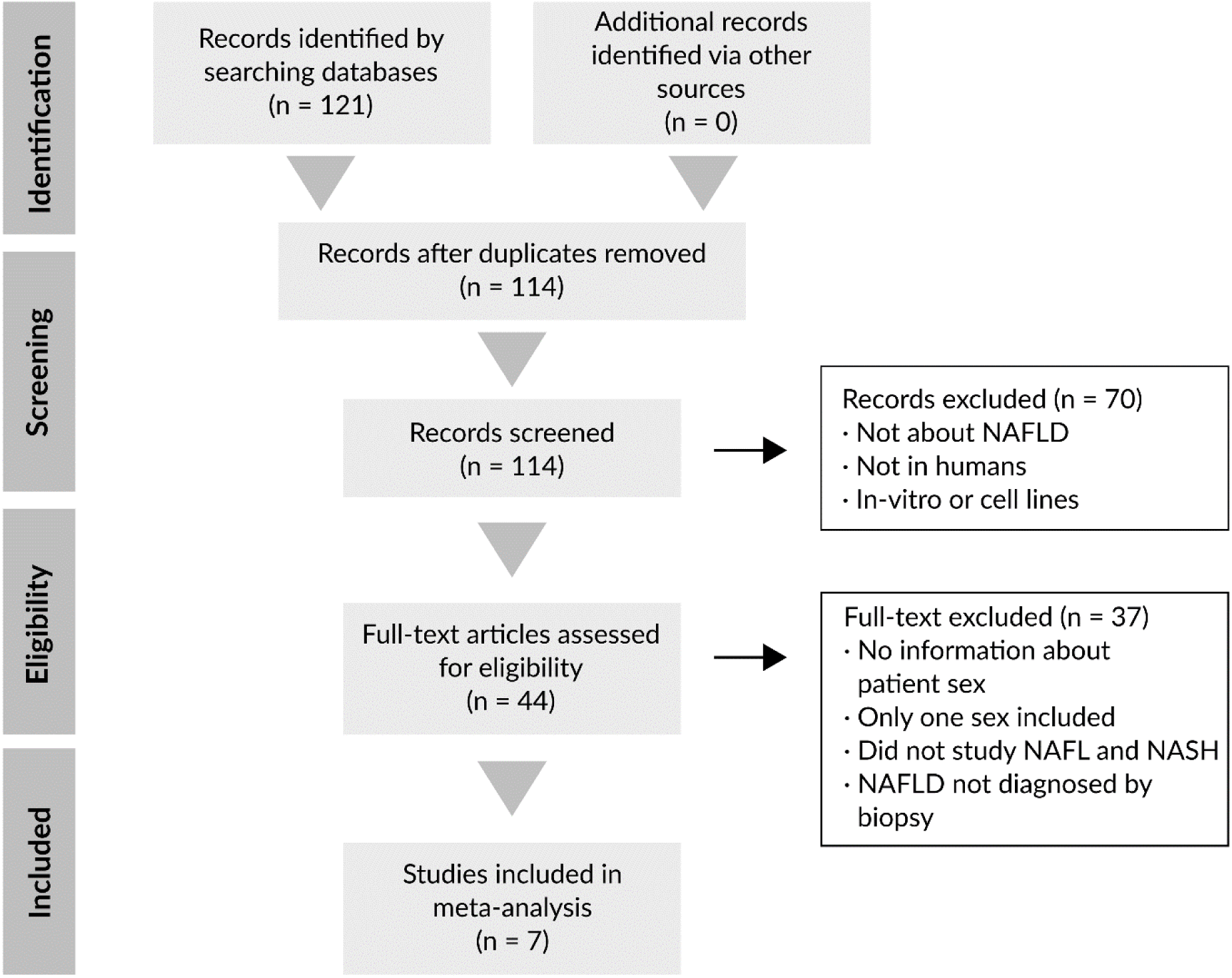
Flow diagram of our systematic review of the literature and selection of studies for this meta-analysis, according to the PRISMA statement guidelines.

The 7 selected studies ^10–16^ included a total of 323 eligible patients (Table 1); 164 individuals were men and 159 women under 50 years (49.2% men and 50.8% respectively). By disease stage, 148 belonged to the NAFL group and 175 to NASH group (45.8 and 54.2%, respectively).

**Table 1.** Characteristics of the studies selected for inclusion in our analysis in terms of disease stage, sex, and patient age.

| GEO<br>accession,<br>Authors, Year<br>[Ref.] | Patients by disease stage and sex |  |  |  | Age |  |
| --- | --- | --- | --- | --- | --- | --- |
|  | Premenopausal<br>women |  | Men |  | Mean | SD |
|  | NAFL | NASH | NAFL | NASH |  |  |
| GSE48452 /<br>Ahrens M. *et al.* 2013 [10] | 10 | 9 | 4 | 4 | 41.62 | 9.35 |
| GSE61260 /<br>Horvath S. *et al.* 2014 [11] | 10 | 7 | 12 | 12 | 41.51 | 9.09 |
| GSE66676 /<br>Xanthakos<br>S.A. *et al.* 2015 [12] | 20 | 4 | 6 | 3 | 17.14 | 1.66 |
| GSE83452 /<br>Lefebvre P. *et al.* 2017 [13] | 32 | 35 | 6 | 46 | 39.61 | 11.51 |
| GSE89632 /<br>Arendt B.M. *et al.* 2015 [14] | 2 | 4 | 14 | 9 | 39.55 | 8.19 |
| GSE126848 /<br>Suppli M.P. *et al.* 2019 [15] | 6 | 4 | 9 | 12 | 40* | 13* |
| GSE130970 /<br>Hoang S.A. *et al.* 2019 [16] | 4 | 12 | 13 | 14 | 44.72 | 9.80 |

\*The information corresponding to the patients in this study was kindly provided by the author of the study, although individual patient ages were not available.

Of note, although the GSE83452 study contained patients at baseline and at one-year post-treatment, only baseline individuals were used in this current study. Table 1 shows the number of individuals from each study that were eligible in our work based on the criteria we established. Our exploratory analysis showed abnormal behaviour for the GSE83452 study; the PCA showed two different blocks: the first 64 samples were grouped into one cluster and the last 88 into another one. We considered this a batch effect and took it into account in our subsequent analyses.

### Individual analysis

The functional analysis of individual genes highlighted significant results overrepresented in both the groups of premenopausal women and of men, as summarised in Table 2. Analysis (using UpSet plots) of the terms or pathways that were common through the different studies showed that none of the elements were common to more than 4 studies (Supplementary Figure 1); in fact, the results for some elements were opposite in different studies (Figure 2A). The functional analysis of individual pathways only exhibited significant results for the GSE83452 study (20 sub-paths and 96 GO terms).

**Table 2.**
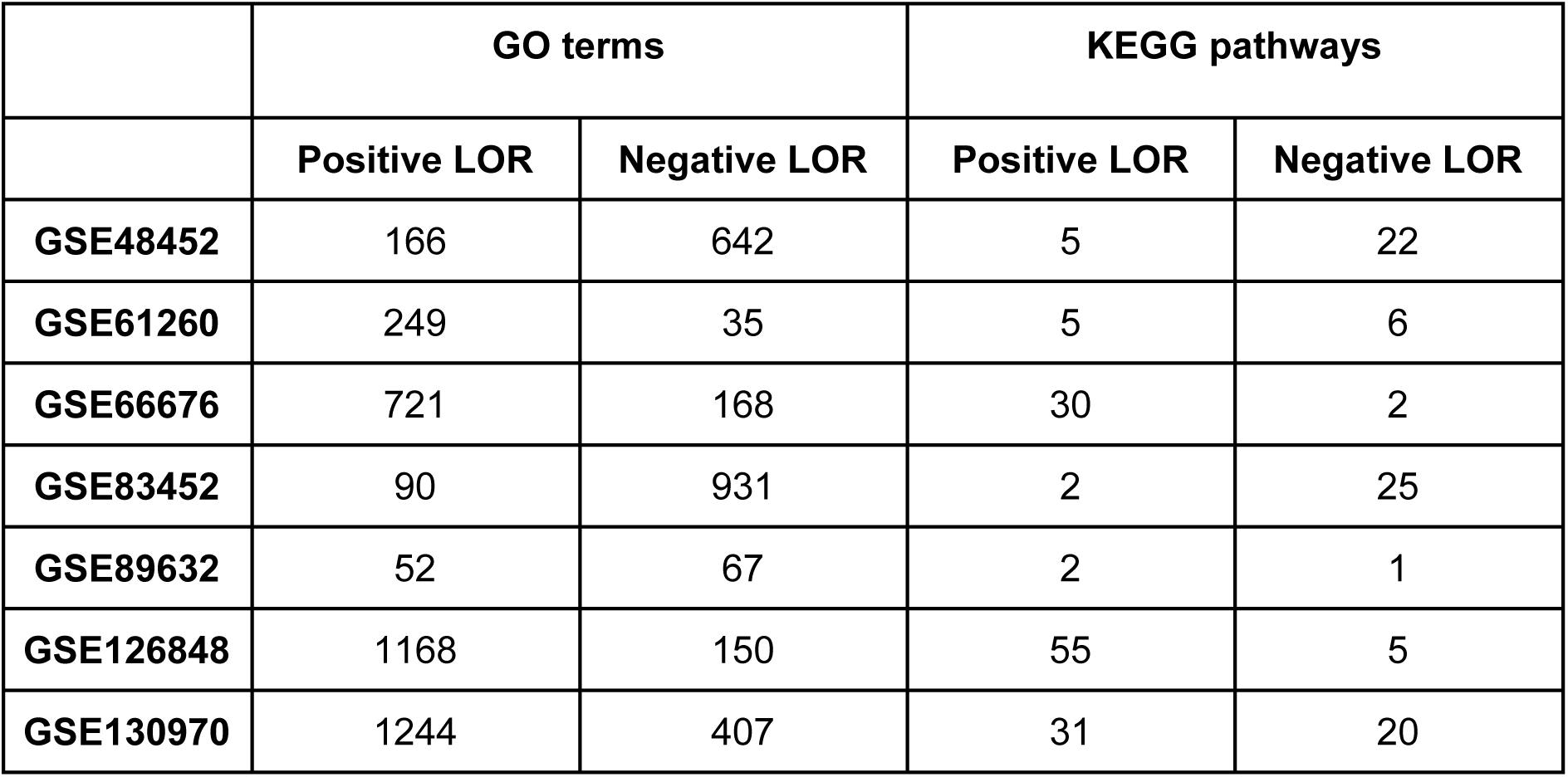
Number of significant GO terms and KEGG pathways in each individual study after gene functional analysis. Positive and negative logarithm of the odds ratio (LORs) represent overrepresentation in premenopausal women and in men, respectively.

|  | GO terms |  | KEGG pathways |  |
| --- | --- | --- | --- | --- |
|  | Positive LOR | Negative LOR | Positive LOR | Negative LOR |
| **GSE48452** | 166 | 642 | 5 | 22 |
| **GSE61260** | 249 | 35 | 5 | 6 |
| **GSE66676** | 721 | 168 | 30 | 2 |
| **GSE83452** | 90 | 931 | 2 | 25 |
| **GSE89632** | 52 | 67 | 2 | 1 |
| **GSE126848** | 1168 | 150 | 55 | 5 |
| **GSE130970** | 1244 | 407 | 31 | 20 |

**Fig. 2.**
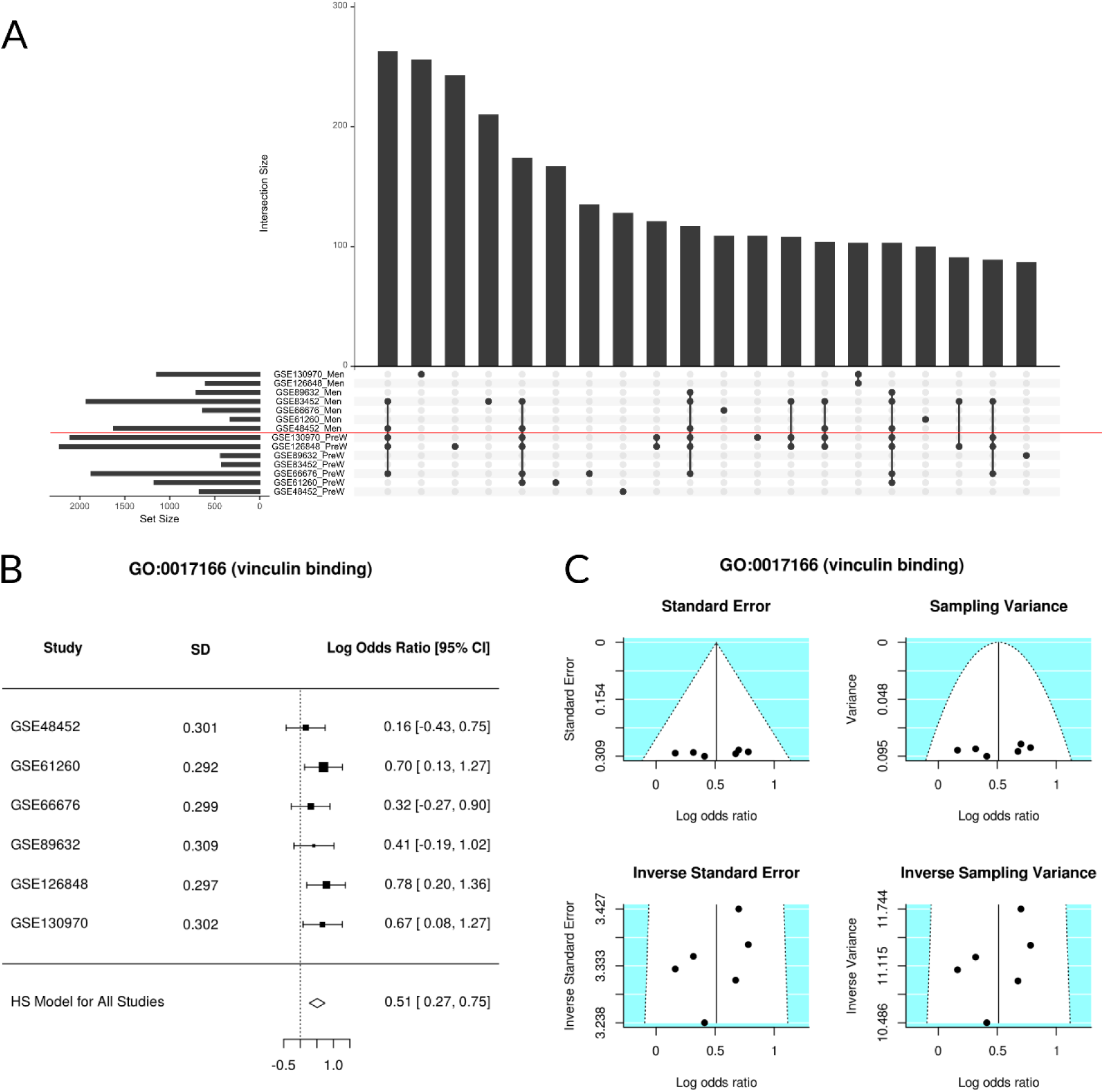
**(A) UpSet plot showing the number of common elements among the significant GO terms in our functional gene analysis**. Only the 20 most abundant interactions are shown. Horizontal bars indicate the number of significant elements in each study; the two groups (premenopausal women and men) are separated by a red line. The vertical bars indicate the common elements in the sets, indicated with dots under each bar. The single points represent the number of unique elements in each group. Cases which cross the red line are significant GO terms, both in premenopausal women and men. **(B) A forest plot of the GO:0017166 term, showing the LOR of each study and the global result. (C) Funnel plot of the GO:0017166 term; dots in the white area indicate the absence of bias and heterogeneity**.

### Meta-analysis

In addition to the *p*-value associated with each of the GO terms, the results of the meta-analysis measured the overall effect of that term throughout all the individual studies, i.e., the LOR. This effect indicated the magnitude of the overexpression of each term (the LOR value) and its direction (the LOR sign). Figure 2B shows, for one function, the LOR of each individual study and the joint LOR of the meta-analysis. The results of all significant elements are detailed in the Supplementary Figures 2 and 3. We confirmed the absence of bias and heterogeneity in all cases, as in Figure 2C (Supplementary Figures 4 and 5).

### Gene functional analysis

The functional meta-analyses of the individual gene differential-expression pipeline results showed a total of 9 significant GO terms (Table 3 and Figure 3), although no KEGG pathways were significant. Among the significant GO terms, 6 were overrepresented in premenopausal women; these elements encompassed a broad spectrum of biological processes and molecular functions, including *RNA polymerase II regulatory region DNA binding* (GO:0001012), *RNA polymerase II regulatory region sequence-specific DNA binding* (GO:0000977), *Vinculin binding* (GO:0017166), *Cellular response to interleukin-2* (GO:0071352), *Negative regulation of neuron death* (GO:1901215), and *Response to interleukin-2* (GO:0070669). The 3 remaining terms were overrepresented in men: *Negative regulation of interleukin-6 secretion* (GO:1900165), *Establishment of planar polarity involved in neural tube closure* (GO:0090177), and *Regulation of establishment of planar polarity involved in neural tube closure* (GO:0090178).

**Table 3.** Significant functions in the functional meta-analyses. The logarithm of the odds ratio (LOR), its confidence interval (LOR-CI), the adjusted *p*-value, and the group in which the function was overexpressed are shown. (W: women; M: men).

|  | GO ID | Name | LOR | LOR-CI | adj. *p*-value | Group |
| --- | --- | --- | --- | --- | --- | --- |
| Gene functional meta-analysis | GO:0001012 | RNA polymerase II regulatory region DNA binding | 0.132 | [0.079, 0.185] | 0.008 | W |
|  | GO:1900165 | Negative regulation of interleukin-6 secretion | -0.762 | [-1.088, -0.437] | 0.018 | M |
|  | GO:0000977 | RNA polymerase II regulatory region sequence-specific DNA binding | 0.131 | [0.073, 0.188] | 0.02 | W |
|  | GO:0017166 | Vinculin binding | 0.511 | [0.271, 0.75] | 0.035 | W |
|  | GO:0071352 | Cellular response to interleukin-2 | 0.467 | [0.248, 0.687] | 0.035 | W |
|  | GO:0090177 | Establishment of planar polarity involved in neural tube closure | -0.514 | [-0.748, -0.279] | 0.035 | M |
|  | GO:0090178 | Regulation of establishment of planar polarity involved in neural tube closure | -0.523 | [-0.768, -0.279] | 0.035 | M |
|  | GO:1901215 | Negative regulation of neuron death | 0.116 | [0.06, 0.171] | 0.043 | W |
|  | GO:0070669 | Response to interleukin-2 | 0.446 | [0.23, 0.662] | 0.046 | W |
| Pathway functional meta-analysis | GO:0098655 | Cation transmembrane transport | 0.194 | [0.11, 0.279] | 0.012 | W |
|  | GO:0030001 | Metal ion transport | 0.141 | [0.074, 0.208] | 0.027 | W |
|  | GO:1904062 | Regulation of cation transmembrane transport | 0.216 | [0.119, 0.32] | 0.027 | W |
|  | GO:0098662 | Inorganic cation transmembrane transport | 0.186 | [0.093, 0.278] | 0.038 | W |

**Fig. 3.**
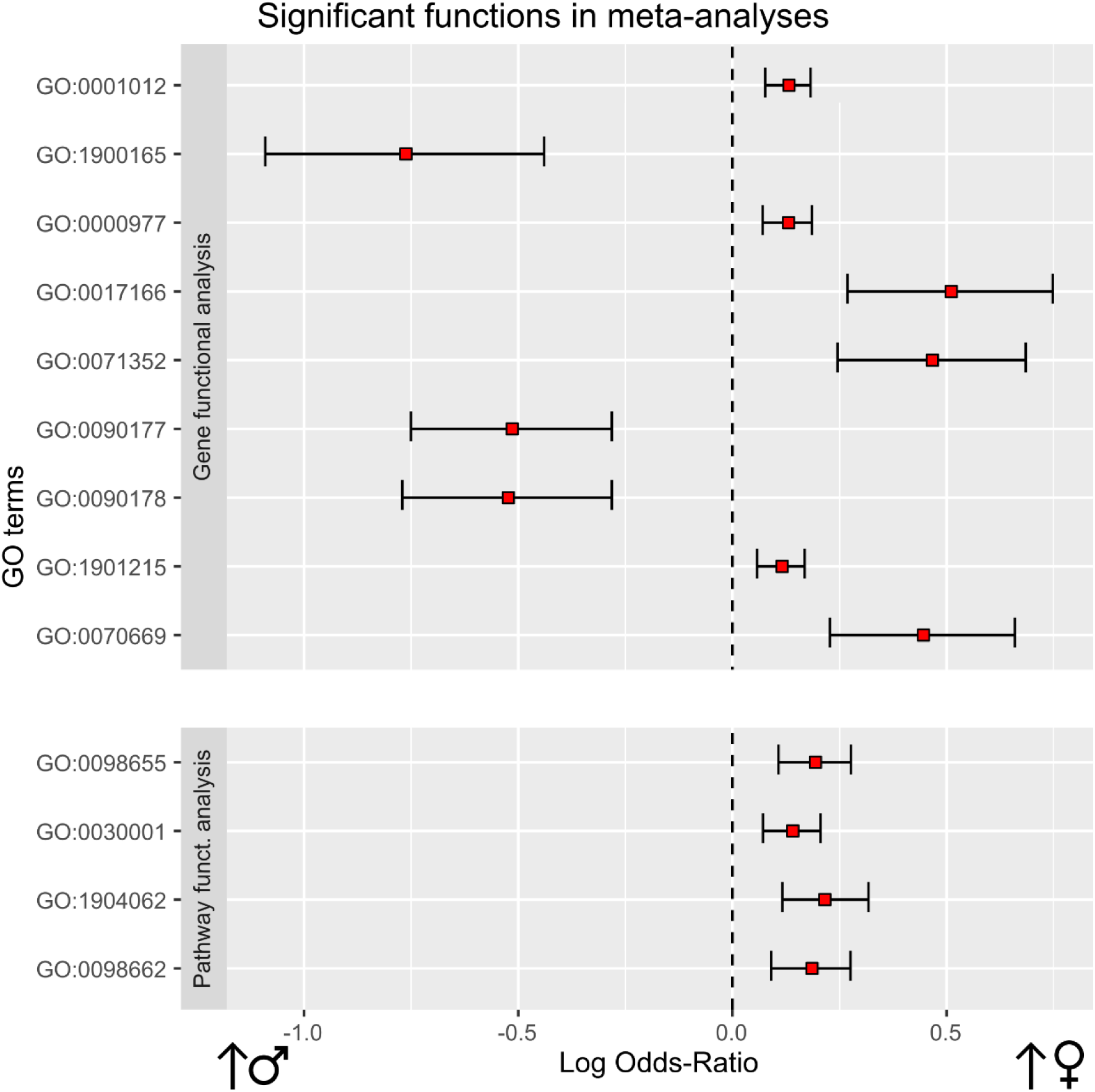
Differential functional profiling by sex. This plot shows significant functional terms of each meta-analysis. On the right: biological functions more overrepresented in women. On the left: biological functions more overrepresented in men. For each function, LOR (red square) and its confidence interval have been represented.

### Pathway functional analysis

The functional meta-analysis of the individual results in the pathway differential-activation pipeline returned a total of 4 significant GO terms, all of them overrepresented in NAFL when comparing with NASH, in premenopausal women and with a LOR value lower than 0.25 (Table 3 and Figure 3). All these terms were related to the transport of ions and cations: *Cation transmembrane transport* (GO:0098655), *Metal ion transport* (GO:0030001), *Regulation of cation transmembrane transport* (GO:1904062), and *Inorganic cation transmembrane transport* (GO:0098662). The meta-analysis of the UniProt functions did not provide any significant results.

### Metafun-NAFLD web tool

Metafun-NAFLD web tool (https://bioinfo.cipf.es/metafun-NAFLD) contains information related to 7 studies and 323 samples used in this work. For each study, the portal included fold-changes of genes and log odds ratios of functions and pathways, which can be explored by users to identify profiles of interest to them.

A total of 8,223 meta-analysis were carried out. For each of the 13 significant functions, metafun-NAFLD showed the global activation level by sex, for all studies and the specific contribution of each one of them, using statistical indicators (log odds ratio, confidence interval and p-value) and graphical representations by function, as forest and funnel plots. This open resource contributes to share data and results between researchers to improve other further works.

## Discussion

Although previous studies have analysed pathobiological mechanisms in NAFLD, and separately sexual differences in metabolic regulation, the impact of gender (sociocultural factors) and sex (biological factors) on NAFLD pathobiology remains incompletely defined. Epidemiological studies have shown a higher prevalence of NAFLD in men and postmenopausal women compared to premenopausal women ^3^. To better understand the molecular basis of these sex differences, here we present a systematic review and functional meta-analysis of publicly available genomic studies, to analyse differences in NAFLD stages, between premenopausal women and men.

Despite the limitations of our approach (presence of studies with different sample sizes and types of platforms), meta-analysis techniques have the capacity to integrate selected studies. The individual results of the 7 selected studies revealed that no GO or KEGG terms were consistently different between groups across all studies, perhaps related to smaller sample sizes, underlying genetic and environmental variance in the source population, and methodological heterogeneity. Also, as shown in Figure 2A, surprisingly, some functions were overexpressed in both groups, depending on the study. Therefore, a meta-analysis, such as this one, can help improve identification of relevant and consistent differences across studies, subject to the caveat that patients in each study may have other differences in clinical characteristics. Also, a meta-analysis can increase the overall sample size and hence the relevance of findings in a disease known to be heterogeneous. In fact, systematic review and meta-analysis is a widely used strategy in the search for sex differences based on available information ^17–19^.

Our functional meta-analysis identified a total of 13 GO terms that were different between men and premenopausal women using two different approaches– differentially expressed genes (9 terms) and differentially activated pathways (4 terms). These terms were related to processes such as inflammation, cell binding, and the establishment of polarity during neural tube closure.

Functional meta-analysis of differentially expressed genes identified differences in 9 significant GO terms. Among these, our results showed the overrepresentation of *Vinculin binding term* (GO:0017166) in premenopausal women compared to men. This cytoplasmic actin-binding protein is enriched in focal adhesions and adherens junctions, contributes to junction stability ^20^, and plays an important role as a regulator of apoptosis ^21^. Of note, although cellular adhesion is a broad topic and not a specific biologic pathway, it was reported to be a GO term consistently enriched in male NASH ^22^ and Arendt et al. also identified genes related to cell-cell adhesion (*ANXA2* and *MAG*) that were differentially expressed between patients with NAFL and NASH ^14^.

Our analysis also identified differences between the sexes related to the inflammatory response, specifically the expression of cytokine-related terms. *Negative regulation of interleukin-6 secretion* (GO:1900165) was overrepresented in men, while *Cellular response to interleukin-2* (GO:0071352) and *Response to interleukin-2* (GO:0070669) were overrepresented in premenopausal women. These terms related to interleukin 2 (IL-2) and interleukin 6 (IL-6), had LOR values close to or above 0.5, indicating a strong difference between the groups. IL-2 induced T cell responses are regulated by the level of IL-2, such that low level IL-2 regulates T-cell central tolerance via the formation of regulatory T cells ^23^. Higher levels of IL-2 induce proliferation of T cells and aid differentiation into memory T-cells. IL-2 receptors are also expressed by B-cells and cells of the innate immune system ^24^. Interestingly, IL-2 signalling is deficient in many chronic liver diseases ^25^ and levels of soluble IL-2 receptor alpha were higher in children with NASH and advanced fibrosis ^26^. These data suggest that low-level IL-2 induced immune tolerance may mitigate the risk of developing NASH in premenopausal women and raise the intriguing possibility of restoring low-level IL-2 signalling in men as a potential treatment of NAFLD.

IL-6 has been previously implicated in NASH with an increased hepatic expression correlating with disease severity as well as insulin resistance ^27^. It is also a regulator of hepatic regeneration ^28^ and a major driver of hepatic carcinogenesis ^29^. In NAFLD, IL-6 may enhance hepatic repair and regeneration, however, it might also be involved in inducing insulin resistance and stimulating hepatocyte apoptosis, thus contributing to NASH transition^28,30^. Indeed IL-6 gene polymorphisms have been associated with an increased susceptibility to chronic liver disease ^31^.

Thus, differences in immune mechanisms between men and women may be one possible factor in for the sex-differences observed in NAFLD. For example, in mice, the macrophage phenotype in males is pro-inflammatory and pro-fibrotic, compared to pro-resolution and anti-fibrotic in women ^32^. To speculate, women may have a relatively decreased immune response to lipotoxicity, whereas, men may have an impaired liver-healing response to chronic injury perhaps related to deficient IL-6 signalling. These ideas remain unsubstantiated and will require further examination and validation in larger studies.

A recent study by Herrera-Marcos et al. detected sex differences in the control of the *Cidec/Fsp27β* gene at the level of the liver ^33^. In fact, expression of the mRNA of this gene was associated with the presence and density of liver lipid droplets in mice fed a high fat diet. Among other functions, *Cidec/Fsp27β* is suggested to be activator for the neuronal pathway in the steatoic liver ^34^. Therefore, the fact that the result for the function *Negative regulation of neuron death* (GO:1901215) was significant in our study could be related to the differences detected by Herrera-Marcos et al.

Interestingly, we found other significant GO terms that had not been associated with NAFLD in previous studies. For instance, *Establishment of planar polarity involved in neural tube closure* (GO:0090177) and *Regulation of establishment of planar polarity involved in neural tube closure* (GO:0090178) were overrepresented in men with a LOR exceeding 0.5 (−0,514 and −0,523). However, we did not find a relationship between these terms and NAFLD.

Our functional pathway meta-analysis identified 4 significant GO terms involved in cation transport: *Cation transmembrane transport* (GO:0098655), *Metal ion transport* (GO:0030001), *Regulation of cation transmembrane transport* (GO:1904062), and *Inorganic cation transmembrane transport* (GO:0098662). The alteration of pathways and functions related to cation transport have been described to be associated with NAFLD ^35^, but information about their specific relevance and significance remains scarce.

Therefore, our results support the notion that there are differences between men and premenopausal women in some biological processes and molecular functions that may be relevant in the NAFLD. Given the well-known heterogeneity in NAFLD progression between males and females this transcriptomic meta-analysis also suggests that pathophysiologically-informed disease biomarkers for males and females may need to be different. Can we further extrapolate these findings to disease definition and clinical trials? For example, biological sex-driven differences may be utilized to better classify any individual with NAFLD and their inclusion into clinical trials. Increased understanding of the differences in the risk of NASH and hepatocellular carcinoma may be included in future practice guidelines. Thus, our meta-analysis underscores the importance of recognizing and accounting for sex-related differences in the study of NAFLD pathobiology.

## Conclusions

Differences in clinical characteristics and pathogenic mechanisms of NAFLD in men versus women have been explored previously but have been not been well characterized. In this regard, starting from a total of 114 GEO studies we selected 7 studies suitable for a functional meta-analysis. We identified a total of 13 significant GO terms from these data, some of which have been previously identified to be relevant to the disease. This supports the notion that sex is an important variable in understanding NAFLD pathobiology and heterogeneity. However, other functional terms we identified have not been previously linked to NAFLD, opening new avenues for further study. In conclusion, meta-analysis of transcriptomic data is a helpful approach to identify sex-related differences between NAFL and NASH.

## Data Availability

This work uses data published in previous studies, which is available in the Gene Expression Omnibus repository.

https://www.ncbi.nlm.nih.gov/geo/query/acc.cgi?acc=GSE48452

https://www.ncbi.nlm.nih.gov/geo/query/acc.cgi?acc=GSE61260

https://www.ncbi.nlm.nih.gov/geo/query/acc.cgi?acc=GSE66676

https://www.ncbi.nlm.nih.gov/geo/query/acc.cgi?acc=GSE83452

https://www.ncbi.nlm.nih.gov/geo/query/acc.cgi?acc=GSE89632

https://www.ncbi.nlm.nih.gov/geo/query/acc.cgi?acc=GSE126848

https://www.ncbi.nlm.nih.gov/geo/query/acc.cgi?acc=GSE130970

http://bioinfo.cipf.es/metafun-NAFLD/

## List of Abbreviations

BH: Benjamini & Hochberg p-value adjusting method
GEO: Gene Expression Omnibus
GO: Gene Ontology
GSA: Gene set analysis
KEGG: Kyoto Encyclopaedia of Genes and Genomes
LOR: Log-odds ratio
NAFLD: Non-alcoholic fatty liver disease
NAFL: Non-alcoholic steatosis
NASH: Non-alcoholic steatohepatitis
PCA: Principal component analysis
PPA: Pathway activity analysis
PRISMA: Preferred Reporting Items for Systematic Reviews and Meta-Analyses
RMA: Robust Multichip Average

## Acknowledgements

The authors would like to thank the Principe Felipe Research Centre (CIPF) for providing access to its computational cluster in order to carry out this work. We would also like to thank Dr. Kristoffer T.G. Rigbolt for providing information about ages of the sample used in study GSE126848.

## Materials and methods

### Search studies

This review was conducted in June 2020, following the Preferred Reporting Items for Systematic Reviews and Meta-Analyses (PRISMA) statement guidelines ^36^. We searched the GEO datasets database ^9^ using the keywords: “NAFLD”, “NAFL”, “steatosis”, “NASH”, and “steatohepatitis” for transcriptomic studies published in English.

### Study exclusion criteria

We applied the following exclusion criteria: (i) studies conducted in organisms other than humans; (ii) studies without information about the sex of the participants or that had not included both sexes; (iii) studies without individuals from the NAFL and NASH stages of the disease; (iv) studies in which the disease had not been diagnosed with a biopsy. In the latter case, we decided to require the use of a biopsy as the method for NAFLD and NASH diagnosis because other less invasive methods such as conventional imaging techniques have a low sensitivity to detect mild steatosis and/or to differentiate NAFL from NASH ^7^.

### Bioinformatics analysis strategy

We applied the same three-step transcriptomic analysis strategy to each separate study: (i) data acquisition and preprocessing; (ii) differential gene expression and functional enrichment analysis; (iii) differential pathway activation and functional analysis. The functional results of all the studies were then integrated using meta-analysis techniques (Figure 4). This work was carried out using the programming language R 3.6.0 ^37^; information about the packages we used and their version numbers are provided in the supplementary material.

**Fig. 4.**
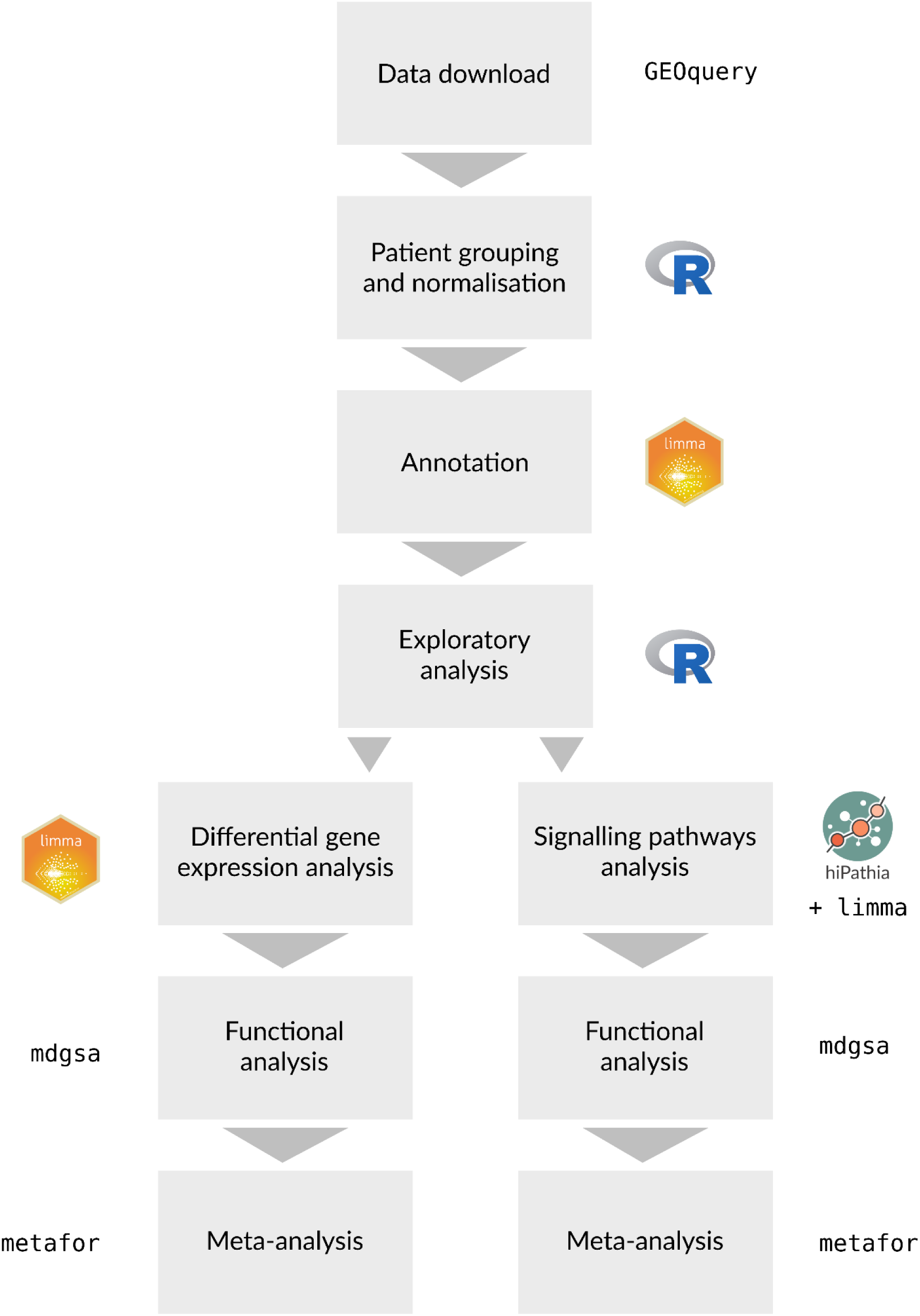
Data-analysis workflow.

### Data acquisition and preprocessing

During the data preprocessing we standardised the nomenclature of the different stages of the disease (by grouping the patients into Control, NAFL, and NASH groups) as well as the association of the probe identifiers with their corresponding genes. In those studies, with treated and untreated patients, treated patients were eliminated for further analysis. For studies that had used microarrays, the probe codes were transformed to their respective Entrez identifiers from the NCBI database.

For repeated probes, we calculated the median of their expression values. When available, data normalised by the original authors of the studies were used. Otherwise (studies GSE61260 and GSE66676), the raw data was normalised using the Robust Multichip Average (RMA) algorithm ^38^. The gene nomenclature in RNA-Seq studies was also standardised to the Entrez identifiers and the raw data matrix was processed using the TMM standardisation method ^39^ followed by a log-transformation of the data.

To meet the objective of this work, women were separated into premenopausal and postmenopausal groups. Since the selected studies did not include this information, we assumed that women aged under 50 years had been premenopausal, based on previous literature indicating that the average age of menopause is around 48–52 years ^40,41^.

After the normalisation of the data we attempted to detect any possible anomalous effects within the studies by completing an exploratory clustering and a principal component analysis (PCA).

### Differential gene expression and functional enrichment analysis

We carried out a differential expression analysis of these studies using the *limma* and *edgeR* packages ^42,43^ to detect genes that were differentially expressed when comparing NAFL and NASH, in premenopausal women versus men. For each gene, we adjusted a linear model, which included possible batch effects, contrasting (NASH.W – NAFL.W) and (NASH.M – NAFL.M), where NASH.W, NAFL.W, NASH.M, and NAFL.M corresponded to NASH-affected premenopausal women, NAFL-affected premenopausal women, NASH-affected men, and NAFL-affected men, respectively. The statistics of the differential expression were calculated and the *p*-values were adjusted using the Benjamini & Hochberg (BH) method ^44^.

Next, based on the differential gene expression results, we performed a functional enrichment analysis using gene set analysis (GSA) ^45^. First, the genes were ordered according to their *p*-value and the sign of the contrast statistic. Next, the GSA was performed using the logistic regression model implemented in the *mdgsa* R package ^46^ along with their corresponding functional annotations obtained from the Gene Ontology (GO) ^47^ and Kyoto Encyclopaedia of Genes and Genomes (KEGG) PATHWAY ^48^ databases.

Because of their hierarchical structure, the gene annotations with GO terms applied in the *mdgsa* package were propagated so that they inherited the annotations of the ancestor terms. Excessively specific or generic annotations (blocks smaller than 10 or larger than 500 words) were subsequently filtered out. Finally, functions with a BH-adjusted *p*-value under 0.05 were considered significant.

For the two functional elements (GO terms and KEGG paths), we analysed the number of over-represented elements shared by the studies. These results were graphically represented as UpSet plots ^49^ to show the number of elements in common between the different sets. First, we compared the overrepresented elements as separate graphs to detect the common functions in each group: GO terms in premenopausal women, GO terms in men, KEGG pathways in premenopausal women, and KEGG pathways in men. We then visualised any significant GO terms in each of the individual studies in the same plot to highlight significant terms with different signs from among the studies.

### Differential pathway activation and functional analysis

The Hipathia algorithm ^50^ was used to perform Pathway Activity Analysis (PAA). This method transforms gene expression values for stimulus-response signalling sub-pathways into the activation levels which ultimately trigger cellular responses. In this current work we used the *hipathia* R package to analyse 1,654 GO terms and 142 UniProt functions ^51^, associated with 146 KEGG routes ^48^.

The activation signal of each subpathway was computed from gene expression values using this algorithm. These values were used to detect significant differential activations in the (NASH.W – NAFL.W) – (NASH.M – NAFL.M) contrast pair (as defined above). The functional annotations included in Hipathia and the differential activation results were integrated using the GSA method to identify differences at the functional level (GO terms and UniProt functions) in relation to NAFLD stages.

### Meta-analysis

Once the gene functional analysis was applied to each individual study, we carried out a functional meta-analysis to summarise the results. Similarly, once the pathway functional analysis was applied to each individual study, the results were summarised via a homologous functional meta-analysis.

Both meta-analyses were performed following the methodology described in detail in García-García ^52^. Briefly, the *metafor* R package ^53^ was used to evaluate the combined effect with a random-effects model; this model more precisely detects overrepresented elements than performing the studies individually at a higher statistical power. Likewise, variability in the individual studies was considered into the global estimation of the measured effect so that less-variable results had a higher weight in the overall calculation of the logarithm of the odds ratio (LOR) ^54^. The incorporation of the variability between experiments, in the model of random effects, provides more statistically robust results and a better integration of selected studies. Finally, we confirmed the suitability of this selection by evaluating the heterogeneity of each study in the global model and the application of cross validation techniques.

For each meta-analysis and function, *p*-values (adjusted using the BH method), LORs, and 95% confidence intervals (CIs) were calculated. A term was considered significant when its adjusted *p*-value was lower than 0.05; a positive LOR in significant functions indicated a greater overrepresentation in premenopausal women than in men. In contrast, a negative LOR indicated higher representation in men than in premenopausal women. Finally, funnel plots and forest plots were used to assess the variability and to measure the contribution of each study to the meta-analysis.

A total of 8,223 elements were analysed in the gene functional meta-analysis (7,994 GO terms and 229 KEGG pathways); 1,654 GO terms and 142 UniProt functions ^51^ were assessed in the functional pathway meta-analysis in association with 146 KEGG pathways ^48^.

The large volume of data and results generated in this work is available in metafun-NAFLD web tool: https://bioinfo.cipf.es/metafun-NAFLD. Its access is free for any user and allows us to check the results shown in this manuscript, as well as to review other results that may be of interest to researchers. The front-end was developed using the Bootstrap library. All the graphics used in this tool have been implemented with Plot.ly except for the exploratory analysis cluster plot which was generated with ggplot2 package.

